# DNA Methylation Signatures of Cardiovascular Health Provide Insights into Diseases

**DOI:** 10.1101/2024.11.19.24317587

**Authors:** Madeleine Carbonneau, Yi Li, Yishu Qu, Yinan Zheng, Alexis C. Wood, Mengyao Wang, Chunyu Liu, Tianxiao Huan, Roby Joehanes, Xiuqing Guo, Jie Yao, Kent D. Taylor, Russell P. Tracy, Durda Peter, Yongmei Liu, W Craig Johnson, Wendy S. Post, Tom Blackwell, Jerome I. Rotter, Stephen S. Rich, Susan Redline, Myriam Fornage, Jun Wang, Hongyan Ning, Lifang Hou, Donald Lloyd-jones, Kendra Ferrier, Yuan-I. Min, April P. Carson, Laura M. Raffield, Alexander Teumer, Hans J. Grabe, Henry Völzke, Matthias Nauck, Marcus Dörr, Arce Domingo-Relloso, Amanda Fretts, Maria Tellez-Plaza, Shelley Cole, Ana Navas-Acien, Meng Wang, Joanne M. Murabito, Nancy L. Heard-Costa, Brenton Prescott, Vanessa Xanthakis, Dariush Mozaffarian, Daniel Levy, Jiantao Ma

**Affiliations:** Population Sciences Branch, Division of Intramural Research, National Heart, Lung, and Blood Institute, National Institutes of Health, Bethesda, MD; Framingham Heart Study, Framingham, MA; Department of Biostatistics, Boston University School of Public Health, Boston, MA; Department of Preventive Medicine, Northwestern University Feinberg School of Medicine, 680 N Lake Shore Drive, Chicago, IL 60611, USA; United States Department of Agriculture (USDA)/ARS Children’s Nutrition Research Center, Baylor College of Medicine, TX, USA; The Institute for Translational Genomics and Population Sciences, Department of Pediatrics, The Lundquist Institute for Biomedical Innovation at Harbor-UCLA Medical Center, 1124 W. Carson Street, Torrance, CA 90502, USA; Department of Pathology & Laboratory Medicine, University of Vermont Larner College of Medicine, 360 South Park Drive, Colchester, VT 05446, USA; Duke Molecular Physiology Institute, Duke University, Durham, NC, USA; Department of Biostatistics, University of Washington, Seattle, WA, USA; Division of Cardiology, Department of Medicine, The Johns Hopkins University School of Medicine, Baltimore, USA; Department of Biostatistics, School of Public Health, University of Michigan, Ann Arbor, MI; Department of Genome Sciences, University of Virginia School of Medicine, 1200 Jefferson Park Avenue, Charlottesville, VA 22903, USA; Division of Sleep and Circadian Disorders, Departments of Medicine and Neurology, Brigham & Women’s Hospital & Harvard Medical School, Boston, MA, 02115, USA; Department of Medicine, Brigham and Women’s Hospital, Boston, MA, USA; Department of Medicine, Harvard Medical School, Boston, MA, USA; Brown Foundation Institute of Molecular Medicine, McGovern Medical School, University of Texas Health Science Center at Houston, 1825 Pressler Street, Houston, TX 77030, USA; Department of Medicine, University of Colorado Anschutz Medical Campus, Aurora, Aurora, CO 80045, USA; Department of Medicine, University of Mississippi Medical Center, 350 W. Woodrow Wilson Avenue, Suite 701, Jackson, MS 39213, USA; Department of Genetics, University of North Carolina at Chapel Hill, 120 Mason Farm Road, Chapel Hill, NC 27599, USA; Department of Psychiatry and Psychotherapy, University Medicine Greifswald, Greifswald, Germany; DZHK (German Centre for Cardiovascular Research), Partner Site Greifswald, Greifswald, Germany; German Centre for Neurodegenerative Diseases (DZNE), Partner Site Rostock/Greifswald, Greifswald, Germany; Department SHIP/Clinical-Epidemiological Research, Institute for Community Medicine, University Medicine Greifswald, Greifswald, Germany; Institute of Clinical Chemistry and Laboratory Medicine, University Medicine Greifswald, Greifswald, Germany; Department of Internal Medicine B, University Medicine Greifswald, Greifswald, Germany; Department of Biostatistics, Columbia University Mailman School of Public Health, New York, NY, USA; Department of Environmental Health Sciences, Columbia University Mailman School of Public Health, New York, NY, USA; Department of Epidemiology, Cardiovascular Health Research Unit, University of Washington, Seattle, Washington, USA; Department of Chronic Diseases Epidemiology, National Center for Epidemiology, Carlos III Health Institute, Madrid, Spain; Population Health Program, Texas Biomedical Research Institute, San Antonio, TX, USA; Nutrition Epidemiology and Data Science, Friedman School of Nutrition Science and Policy, Tufts University, Boston, MA; Department of Medicine, Section of General Internal Medicine Boston University Chobanian & Avedisian School of Medicine, Boston, MA and Boston Medical Center, Boston, MA; Section of Preventive Medicine and Epidemiology, Boston University School of Medicine, Boston, MA

## Abstract

**Background:** The association of overall cardiovascular health (CVH) with changes in DNA methylation (DNAm) has not been well characterized.

**Methods:** We calculated the American Heart Association’s Life’s Essential 8 (LE8) score to reflect CVH in five cohorts with diverse ancestry backgrounds. Epigenome-wide association studies (EWAS) for LE8 score were conducted, followed by bioinformatic analyses. DNAm loci significantly associated with LE8 score were used to calculate a CVH DNAm score. We examined the association of the CVH DNAm score with incident CVD, CVD-specific mortality, and all-cause mortality.

**Results:** We identified 609 CpGs associated with LE8 score at false discovery rate (FDR) < 0.05 in the discovery analysis and at Bonferroni corrected *P* < 0.05 in the multi-cohort replication stage. Most had low-to-moderate heterogeneity (414 CpGs [68.0%] with I^2^ < 0.2) in replication analysis. Pathway enrichment analyses and phenome-wide association study (PheWAS) search associated these CpGs with inflammatory or autoimmune phenotypes. We observed enrichment for phenotypes in the EWAS catalog, with 29-fold enrichment for stroke (*P* = 2.4e-15) and 21-fold for ischemic heart disease (*P* = 7.4e-38). Two-sample Mendelian randomization (MR) analysis showed significant association between 141 CpGs and ten phenotypes (261 CpG-phenotype pairs) at FDR < 0.05. For example, hypomethylation at cg20544516 (*MIR33B*; *SREBF1*) associated with lower risk of stroke (*P* = 8.1e-6). In multivariable prospective analyses, the CVH DNAm score was consistently associated with clinical outcomes across participating cohorts, the reduction in risk of incident CVD, CVD mortality, and all-cause mortality per standard deviation increase in the DNAm score ranged from 19% to 32%, 28% to 40%, and 27% to 45%, respectively.

**Conclusions:** We identified new DNAm signatures for CVH across diverse cohorts. Our analyses indicate that immune response-related pathways may be the key mechanism underpinning the association between CVH and clinical outcomes.

## INTRODUCTION

Cardiovascular disease (CVD) remains the most frequent cause of death in the US and globally.^1^ To promote cardiovascular health (CVH) prior to onset of clinical CVD, the American Heart Association (AHA) has developed a new CVH metric, Life’s Essential 8 (LE8),^2^ which includes the four health factors: BMI, blood lipids, blood glucose, and blood pressure and the four health behaviors: dietary habits, physical activity, smoking, and sleep. Several recent studies have shown that a favorable CVH score is associated with lower risk of CVD and mortality.^3-6^ The maximum CVH score is 100; but over 80% of US adults have low (<50, 17.9% of adults) or moderate (50-79, 62.5%) scores of CVH; only 19.6% have ideal CVH (scores of 80-100).^7^ Attaining ideal CVH is challenging, partly because promoting healthy behaviors is not a trivial task, e.g., public health efforts to improve diet quality and physical activity have been met with limited success.^8^ Identifying omics biomarkers of CVH has the potential to uncover key molecular mechanisms and biological pathways linking poor CVH to increased risk of CVD and mortality. With this insight, we may be able to design more precise and effective prevention and intervention strategies to improve CVH.

DNA methylation (DNAm) occurs predominantly on cytosine residues of the dinucleotide sequence (i.e., CpGs).^9, 10^ In large-scale observational studies, DNAm marks are often measured using blood samples because of the minimal invasiveness and accessibility, as well as the critical role of blood-derived DNAm in crosstalk with other tissues. Differential DNAm patterns have been associated with CVH components and CVD risk.^11-17^ Furthermore, genetic instruments (e.g., Mendelian randomization [MR] analysis) suggest that DNAm at several CpGs are causally related to CVD.^16, 18^ These findings suggest that DNAm biomarkers provide promise for better understanding the epigenetic pathways and molecular mechanisms of both CVH and CVD. We have shown that Life’s Simple 7 (LS7), the AHA’s CVH score prior to LE8, was associated with a DNAm signature.^15^ Recently, we also demonstrated that the AHA’s LE8 score was associated with DNAm-based biological age;^19^ and that DNAm age markers may explain up to 40% of the association of the LE8 score with incident CVD.^19^ However, previous studies linking CVH, DNAm, and onset of clinical CVD have been limited by moderate sample size or lack of external validation. To better characterize the DNAm signature of CVH and its clinical implications, we conducted an epigenome-wide association study (EWAS) of CVH with accompanying pathway enrichment analyses, phenome-wide association study (PheWAS), MR analyses, and prospective investigation of incident CVD and mortality across six large and diverse cohorts. We hypothesized that we could identify specific DNAm signatures and corresponding molecular mechanisms underlying the relationship between CVH and CVD and mortality.

## METHODS

### Study populations

This investigation leverages participant-level data from the Framingham Heart Study (FHS),^20^ Coronary Artery Risk Development in Young Adults Study (CARDIA),^21^ Multi-Ethnic Study of Atherosclerosis (MESA),^22^ The Study of Health in Pomerania (SHIP),^23^ Strong Heart Study (SHS),^24, 25^ and Jackson Heart Study (JHS).^26^ Additional information on each cohort is provided in the **Supplemental Materials**. Protocols and procedures for the study cohorts were approved by the Institute Review Board of each cohort. The current study was approved by the Institutional Review Board at Tufts University.

### LE8 score calculation

We calculated the CVH score (i.e., the LE8 score) using the approach proposed by the AHA,^2^ with modifications to accommodate data availability in five cohorts (CARDIA, FHS, MESA, SHIP, and SHS). Procedures used to calculate LE8 scores for each cohort are detailed in **Supplemental Table 1**. Briefly, each component was scored separately from 0 to 100. The LE8 score was calculated as the unweighted mean of all individual component scores. In FHS, CARDIA, and SHIP, the LE8 score was calculated using all eight components, while this score was calculated based on seven components in MESA (no data for sleep health) and six components in SHS (no data for sleep health and diet). We tested and confirmed that the correlation between the LE8 score with all components and the score calculated with missing components was high (**Supplemental Table 2**).

### DNA methylation (DNAm) measurement

DNAm levels were measured based on whole blood samples in all cohorts. Details of DNA collection and assessment of DNAm have been described in the **Supplemental Materials.** Briefly, Illumina MethylationEPIC BeadChip array (EPIC array) was used in all cohorts except FHS, which used the Illumina HumanMethylation450 BeadChip array (450K array). Cohort-specific quality control and normalization had been performed following standard protocols. The 450K array targets 485,512 CpGs and the EPIC array targets 866,836 CpGs; ∼93% of CpGs in the 450K array are covered by the EPIC array.^27^ Because FHS was our largest participating cohort, we primarily analyzed CpGs that are common to both arrays. We also excluded probes on the X and Y chromosomes to avoid potential sex bias. The methylation level at each CpG was quantified as a β value, i.e., the proportion of methylated CpGs over the sum of methylated and unmethylated CpGs.

### Clinical outcome ascertainment

Prospective outcomes analyzed were incident CVD, CVD-specific mortality, and all-cause mortality. In each participating study, these clinical events were adjudicated by physicians using medical and hospital records, death certificates, and next-of-kin interviews.^28^ CVD definition by studies was reported in **Supplemental Table 3**. Participants with CVD identified at baseline (i.e., prevalent CVD cases) were removed before analysis for incident CVD and CVD-specific mortality.

### Statistical analysis

We performed three main analyses (**Figure 1**), including 1) conducting an EWAS and meta-analysis for cross-sectional associations of the LE8 score and DNAm, 2) using various bioinformatic tools to explore potential biological and clinical implications for LE8 score-associated CpGs, and 3) creating a LE8 DNAm score and testing its prospective association with clinical outcomes.

**Figure 1.**
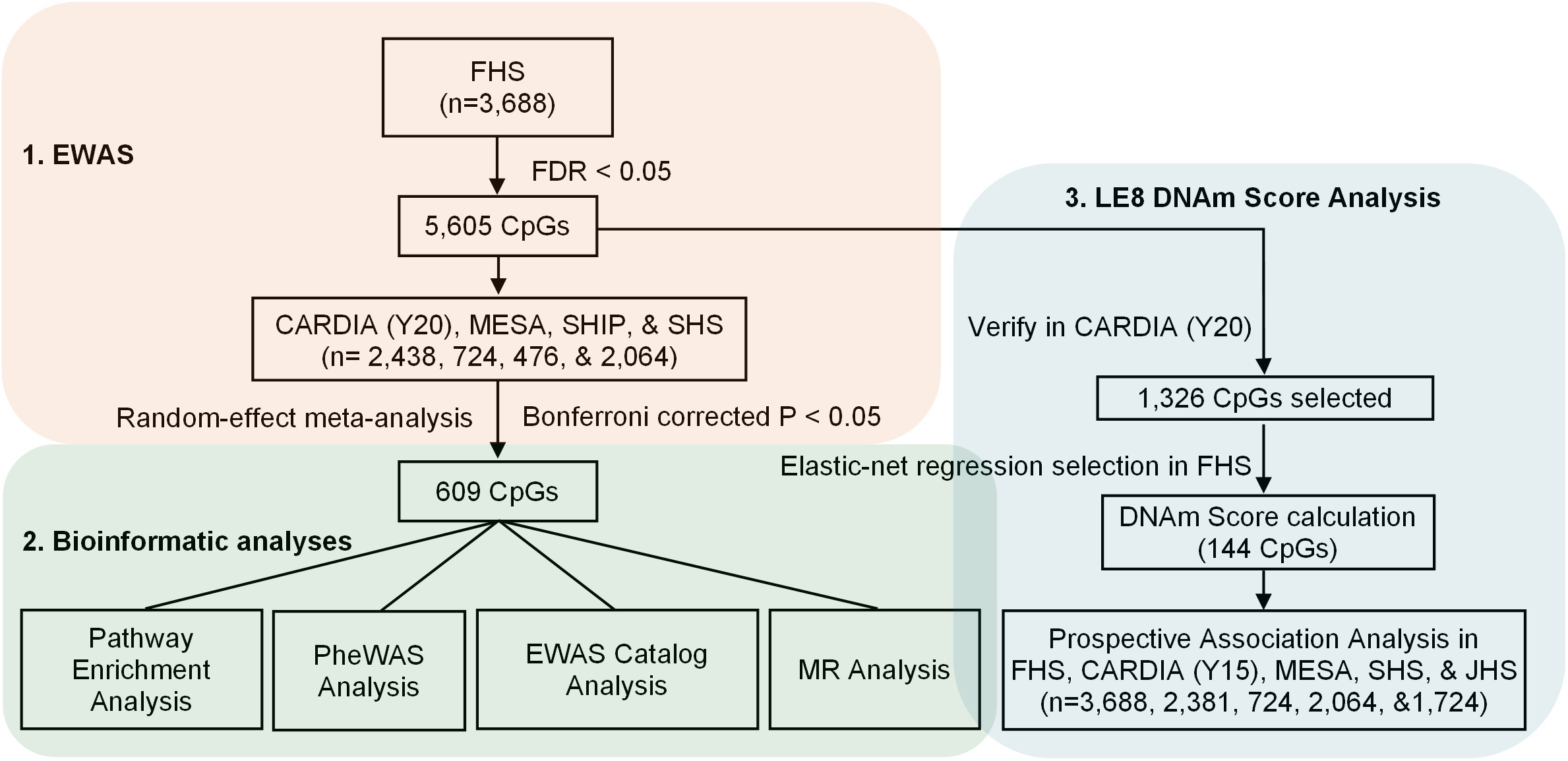
Study design flow-chart. EWAS: epigenome-wide association study, PheWAS: phenome-wide association study, DNAm: DNA methylation, MR: Mendelian randomization, FHS: Framingham Heart Study, CARDIA: Coronary Artery Risk Development in Young Adults Study, MESA: Multi-Ethnic Study of Atherosclerosis, SHIP: The Study of Health in Pomerania, SHS: Strong Heart Study, JHS: Jackson Heart Study

### EWAS of LE8 score and DNAm

Due to its large sample size and homogenous study sample, we first performed discovery EWAS in FHS. For CpGs reaching significance, i.e., those with false discovery rate (FDR) < 0.05, in the discovery analysis, we performed validation analysis using meta-analysis of EWAS results from four independent cohorts (CARDIA, MESA, SHIP, and SHS). In FHS, to remove potential batch effects, DNAm β values for 443,238 autosomal CpGs were residualized by regressing on surrogate variables^29^ using linear regression models.^11^ We then used linear mixed models to examine associations of the LE8 score with DNAm residuals as the dependent variable and LE8 score as the independent variable. Covariates were sex, age, and leukocyte composition estimated using the Houseman method.^29^ Familial relatedness in FHS participants was considered as a random-effect factor. In each replication cohort, we also residualized DNAm β values and performed ancestry-specific LE8 score-CpG associations analysis using linear regression models with adjustment for the same covariates as those included in FHS. In SHS, to achieve more stable inference, an empirical Bayes method (in *limma* R package^30^) was used to moderate the standard errors. In CARDIA, a Bayesian method (in *bacon* R package^31^) was used to adjust the standard errors based on estimation of the empirical null distribution. We performed random-effect meta-analysis to calculate pooled effect size, standard errors, *P* values, and heterogeneity^32^ (*I^2^*) in replication cohorts. We considered CpGs replicable if the association direction was same to those in FHS and meta-analysis *P* reached < 0.05 after a Bonferroni correction by the number of significant CpGs in FHS. All replicated CpGs were annotated to genes using the Illumina Methylation array annotation files.

### Pathway enrichment analyses

We examined whether LE8-associated CpGs were enriched in any biological pathways. In this analysis, we searched gene symbols annotated to the LE8-associated CpGs in two platforms, the Database for Annotation, Visualization and Integrated Discovery (DAVID)^33, 34^ and the Functional Mapping and Annotation of Genome-Wide Association Studies (FUMA)^35^. Multiple gene sets such as the gene ontology (GO)^36^ terms of biological function, Kyoto Encyclopedia of Genes and Genomes (KEGG),^37^ Molecular Signatures Databases (MSigDB),^38^ WikiPathways,^39^ and GWAS catalog reported genes^40^ were examined. Fisher’s exact test was used to calculate enrichment *P* values, and a test with FDR < 0.05 was considered statistically significant. We visualized the networks of enriched pathways (i.e., clusters) using the R package *FGNet*.^41^

### Overlap with EWAS catalog

LE8 and its subcomponents have been suggested to be associated with a wide range of phenotypes across the life course.^2, 42, 43^ We therefore mined EWAS catalog, a database for CpGs associated with various traits (https://www.ewascatalog.org),^17^ for prior evidence of disease related associations for the replicated LE8-associated CpGs. By overlap of these CpGs with those reported in EWAS catalog, we examined whether LE8-associated CpGs were linked with major diseases such as CVD, neurodegenerative diseases, and cancer. We filtered out EWAS catalog CpGs with *P* > 1e-6 and CpGs identified in studies with sample size < 500 individuals in this catalog. Fisher’s exact tests were performed to determine if LE8-associated CpGs were enriched with EWAS catalog CpGs for disease phenotypes. A test with FDR < 0.05 was considered statistically significant.

### Phenome-wide association study (PheWAS) catalog analysis

We further searched the PheWAS catalog (https://phewascatalog.org) for disease phenotypes that may be associated with LE8-associated CpGs. The PheWAS catalog included 1,358 phenotypes based on electronic medical records of 13,835 individuals of European ancestry in the eMERGE Network.^44^ We searched this catalog using DNAm quantitative trait loci (meQTL) single nucleotide polymorphisms (SNPs) in the Genetics of DNA Methylation Consortium (GoDMC) meQTL database (http://mqtldb.godmc.org.uk)^45^ for LE8-associated CpGs. We focused on *cis*-meQTL SNPs (defined as SNPs residing within 1Mb from the corresponding CpG with *P* < 1e-8) in GoDMC database and SNPs that were also reported in the GWAS catalog.^40^ We considered SNPs with *P* < 3.7e-5 (0.05/1,358) significant in the PheWAS catalog. For significant SNPs, we further performed PheWAS search in the IEU OpenGWAS database.^46^ We included ten batches of GWAS summary datasets such as UK Biobank phenotypes for up to 3,873 phenotypes (https://gwas.mrcieu.ac.uk/datasets; **Supplemental Table 4**). We reported SNP-phenotype association with *P* values < 1.3e-5 (0.05/3,837) in the IEU OpenGWAS database.^46^

### Mendelian randomization (MR) analysis

To further explore the relationship between LE8-associated CpGs with CVD and other disease phenotypes, we used the *TwoSampleMR* R package^47^ to test the putative causal association of LE8-associated CpGs with the 14 phenotypes, including stroke, heart failure, atrial fibrillation, coronary arterial calcification (CAC), COPD, C-reactive protein (CRP), % lymphocytes, % monocytes, % neutrophils, preeclampsia, gestational hypertension, Alzheimer’s disease, lung cancer, and breast cancer.^48-56^ We used independent *cis*-meQTL SNPs from the GoDMC database, after linkage disequilibrium (LD) pruning based on R^2^ < 0.01, as instruments. We extracted effect sizes (regression coefficients and standard errors) from both the GoDMC *cis*-meQTL database^57^ and the GWAS summary results for the 14 phenotypes (**Supplemental Table 5**). We performed the primary analysis using the inverse variance weighted (IVW) method and sensitivity analysis using the MR-Egger method when we identified three or more instrumental SNPs. CpGs with phenotype-specific FDR < 0.05 were considered significant.

### CVH DNAm score

We developed the CVH DNAm score based on FHS and CARDIA EWAS results because the LE8 score was calculated using all eight components with large sample size. Also, this design eliminated the potential overfitting in analysis for the association between CVH DNAm score and clinical outcomes in other participating cohorts (MESA, SHS, and JHS). We first selected CpGs with FDR < 0.05 in FHS and Bonferroni corrected *P* values < 0.05 in CARDIA (i.e., corrected by the number of CpGs with FDR < 0.05 in FHS). Using participant-level data from FHS, we performed further CpG selection using elastic-net regression.^58^ For individuals with missing values for certain CpGs, cohort median values were assigned. Notably, the proportion of missingness was very low (0.01%), and the impact of missingness was negligible. Before entering the elastic-net regression model, we calculated CpG residuals using linear regression models with adjustment for sex, age, estimated leukocyte composition, laboratories for DNAm measurements, and DNAm batch variables. To tune the hyperparameter λ, we performed 10-fold cross-validation with default tuning grid (i.e., α = 0.1, 0.55, and 1) using R package *caret*^59^ and *glmnet*.^60^ Elastic-net regression selected CpGs were aggregated into a single sum score, with weights applied reflecting regression coefficients in the meta-analysis of FHS and CARDIA. The same selected CpGs and weights were also applied to derive CVH DNAm scores in other participating studies.

### Prospective association of the CVH DNAm score and clinical outcomes

Cox models were used to examine the prospective association between the CVH DNAm score and incident CVD, CVD-specific mortality, and all-cause mortality in each cohort. Mixed Cox models were used to account for familial relationships in FHS. To reduce potential model overfitting in this prediction analysis in CARDIA, we used DNAm data measured five years (i.e., at Year 15) prior to those used for LE8 EWAS (i.e., at Year 20). Two sets of covariates were examined, including 1) sex, age, estimated leukocyte composition, cohort-specific covariates such as research centers, and self-reported race/ethnicity and 2) model 1 covariates plus education levels. Harrell’s C statistic was calculated to evaluate the model concordance.^61^ The proportional hazards assumption for a Cox regression model fit was examined based on weighted residuals, using the cox.zph function in *survival* R package.^62^ Two-sided *P* values < 0.05 were considered as significant in this analysis.

## RESULTS

### Participant characteristics

**Table 1** displays participant characteristics for all six cohorts included in this study. The study sample for the CVH EWAS and DNAm score analysis included 9,390 and 10,581 participants, respectively. CARDIA participants were generally younger than those in other cohorts. The mean age of CARDIA participants at year 15 and year 20 were ∼38 years and ∼43 years. Mean age ranged 51 to 61 years in the other cohorts. About half (51.3%) of study participants were of European ancestry; and the remainder of Native American (18.6%) and African (27.6%), Hispanic (2%), and Chinese (0.5%) ancestry.

**Table 1:** Participant characteristics by participanting studies.

|  | FHS | CARDIA (Y15) | CARDIA (Y20) | MESA | SHIP | SHS | JHS |
| --- | --- | --- | --- | --- | --- | --- | --- |
| Total Participants (N) | 3688 | 2381 | 2438 | 724 | 476 | 2064 | 1724 |
| Mean Age, years (SD) | 58 (13) | 40 (4) | 45 (4) | 61 (10) | 51 (14) | 56 (8) | 56 (12) |
| Sex, % Female | 2008 (54%) | 1406 (59%) | 1401 (57%) | 332 (46%) | 217 (46%) | 1205 (58%) | 1080 (63%) |
| Smoking status, n (%) |  |  |  |  |  |  |  |
| Never | 2489 (68%) | 1471 (62%) | 1504 (62%) | 402 (56%) | 186 (39%) | 667 (32%) | 1115 (65%) |
| Former | 890 (24%) | 451 (19%) | 502 (21%) | 321 (44%) | 167 (35%) | 606 (29%) | 356 (21%) |
| Current | 309 (8%) | 459 (19%) | 431 (18%) | 1 (0.1%) | 123 (26%) | 791 (38%) | 243 (14%) |
| NA |  |  | 1 (0.04%) |  |  |  | 10 (0.6%) |
| Race/Ethnicity |  |  |  |  |  |  |  |
| AA |  | 987 (41%) | 1068 (44%) | 123 (17%) |  |  | 1724 (100%) |
| EA | 3688 (100%) | 1394 (59%) | 1370 (56%) | 317 (44%) | 476 (100%) |  |  |
| HA |  |  |  | 224 (31%) |  |  |  |
| CHN |  |  |  | 60 (8%) |  |  |  |
| American Indian |  |  |  |  |  | 2064 (100%) |  |
| DASH diet score (or cohort-specific diet quality score) (SD) | 24 (6) |  | 68 (12) * | 41 (11) | 14 (3) |  |  |
| Physical activity score (SD) | 36 (6) | 345 (275) | 341 (280) | 1665 (2423) * | 2 (1) | 2 (2) |  |
| Sleep, hours (SD) | 7.3 (1.1) | 6.5 (1.2) | 6.7 (1.6) |  | 7 (1.2) |  |  |
| BMI, kg/m2 (SD) | 28 (5.4) | 28.6 (6.8) | 29.3 (6.7) | 28.6 (5.1) | 27.3 (4.2) | 29.7 (2) | 32.0 (7.4) |
| Systolic blood pressure, mm Hg (SD) | 123 (17) | 112 (14) | 116 (15) | 123 (20) | 124 (17) | 124 (6) | 128 (16) |
| Diastolic blood pressure, mm Hg, (SD) | 74 (10) | 74 (11) | 73 (11) | 71 (10) | 76 (9) | 76 (4) | 76 (9) |
| Hypertension meds, n (%) | 1326 (36%) | 168 (7%) | 406 (17%) | 253 (35%) | 145 (31%) | 411 (20%) | 934 (54%) |
| Total cholesterol, mg/dL (SD) | 186 (35) | 185 (35) | 185 (35) | 193 (33) | 211 (43) | 193 (13) | 199 (41) |
| Triglyceride, mg/dL (SD) | 115 (72) | 105 (92) | 110 (81) | 126 (63) | 123 (66) | 119 (26) | 107 (70) |
| HDL, mg/dL (SD) | 58 (18) | 51 (14) | 54 (17) | 51 (14) | 57 (14) | 44 (5) | 51 (15) |
| Lipid lowering meds, n (%) | 1255 (34%) | 47 (2%) | 228 (9%) | 122 (17%) | 60 (13%) |  | 237 (14%) |
| Blood glucose, mg/dL (SD) | 102 (21) | 94 (18) | 99 (25) | 94 (23) | 97 (11) | 110 (18) | 91 (9) |
| HbA1c, % (SD) | 5.6 (0.6) |  | 5.5 (0.9) |  | 5.2 (0.5) | 5.4 (0.7) | 6.1 (1.4) |
| Diabetic meds, n (%) | 240 (7%) | 36 (2%) | 109 (5%) | 51 (7%) | 0 (0%) |  | 306 (18%) |

\* In CARDIA, diet was evaluated based on healthy eating index (HEI) at Year 20 exam
\* In MESA, physical activity level: (hours of intense activity or metabolic equivalent (METs) for sedentary, light, moderate, and intense activities)

### EWAS identified 609 LE8-associated CpGs

We performed association analyses between the LE8 score and 443,238 CpGs in FHS. We also calculated the associations and performed meta-analysis for 783,289 CpGs with complete DNAm data in all four replication cohorts (CARDIA, MESA, SHIP, and SHS). QQ plots of FHS EWAS and meta-analysis of the replication cohorts are shown in **Supplemental Figure 1**. Out of the 443,238 CpGs, associations for 397,905 (89.8%) CpGs were included in the validation analysis. At FDR < 0.05, we observed that the LE8 score was associated with DNAm levels at 5,605 CpGs in FHS, and of these, 5,223 (83.2%) were available in the replication cohort meta-analysis. After Bonferroni correction for 5,223 tests, 609 CpGs were statistically significant (i.e., *P* < 0.05/5,223) in the replication cohorts (**Supplemental Table 6**). For each of these 609 CpGs, the direction of the association in FHS was the same as that in the replication cohorts. In the combined meta-analysis of all five cohorts (783,289 CpGs; **Figure 2**), the majority of the 609 CpGs had *P* < 0.05/783,289 (501 CpGs [82.3%]) and low to moderate heterogeneity (553 CpGs [90.8%] with I^2^ < 0.5; 414 CpGs [68.0%] with I^2^ < 0.2).

**Figure 2.**
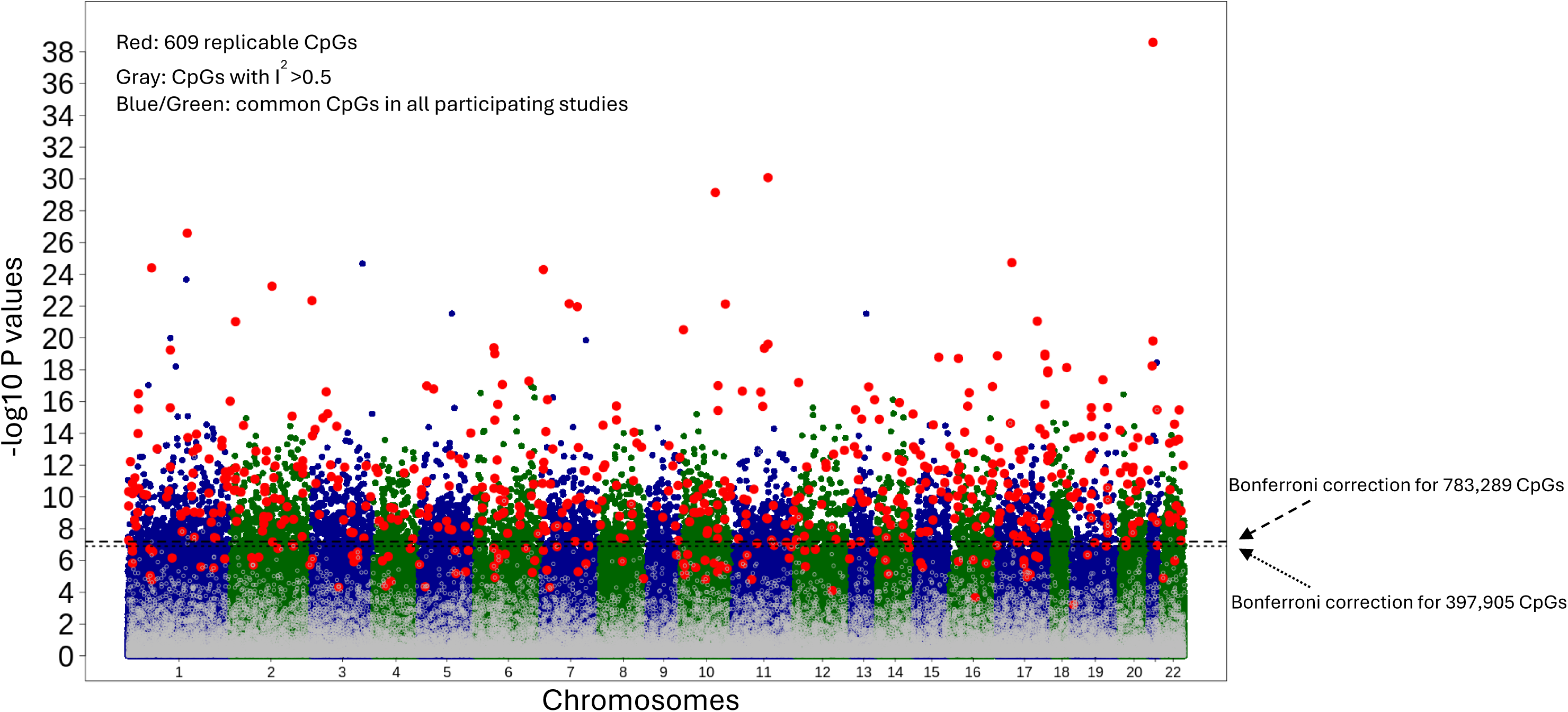
Manhattan plot for LE8 EWAS. Plot was generated using data from combined analysis of all five cohorts (FHS, CARDIA, MESA, SHIP, and SHS). The number of common CpGs in all five cohorts was 397,905 and the number of common CpGs in replication cohorts were 783,289.

### Enrichment analysis demonstrated multiple biological pathways

We observed enrichment for 80 GO terms involved with biological processes (level 3) at FDR < 0.05. The top three significant GO terms were single-organism transport (GO:0044765; *P* = 1.2E-07; FDR = 2.9E-05), cellular response to chemical stimulus (GO:0070887; *P* = 1.4E-07; FDR = 2.9E-05), and regulation of cell communication (GO:0023051; *P* = 1.4E-07; FDR = 2.9E-05). Our GO network analysis further showed clustering of the GO terms (**Figure 3**). We observed nine clusters for 76 GO terms (**Supplemental Table 7**), e.g., those related to cell communication and signaling (Cluster 2), cellular immune response (Cluster 5), and development and cellular differentiation (Cluster 3).

**Figure 3.**
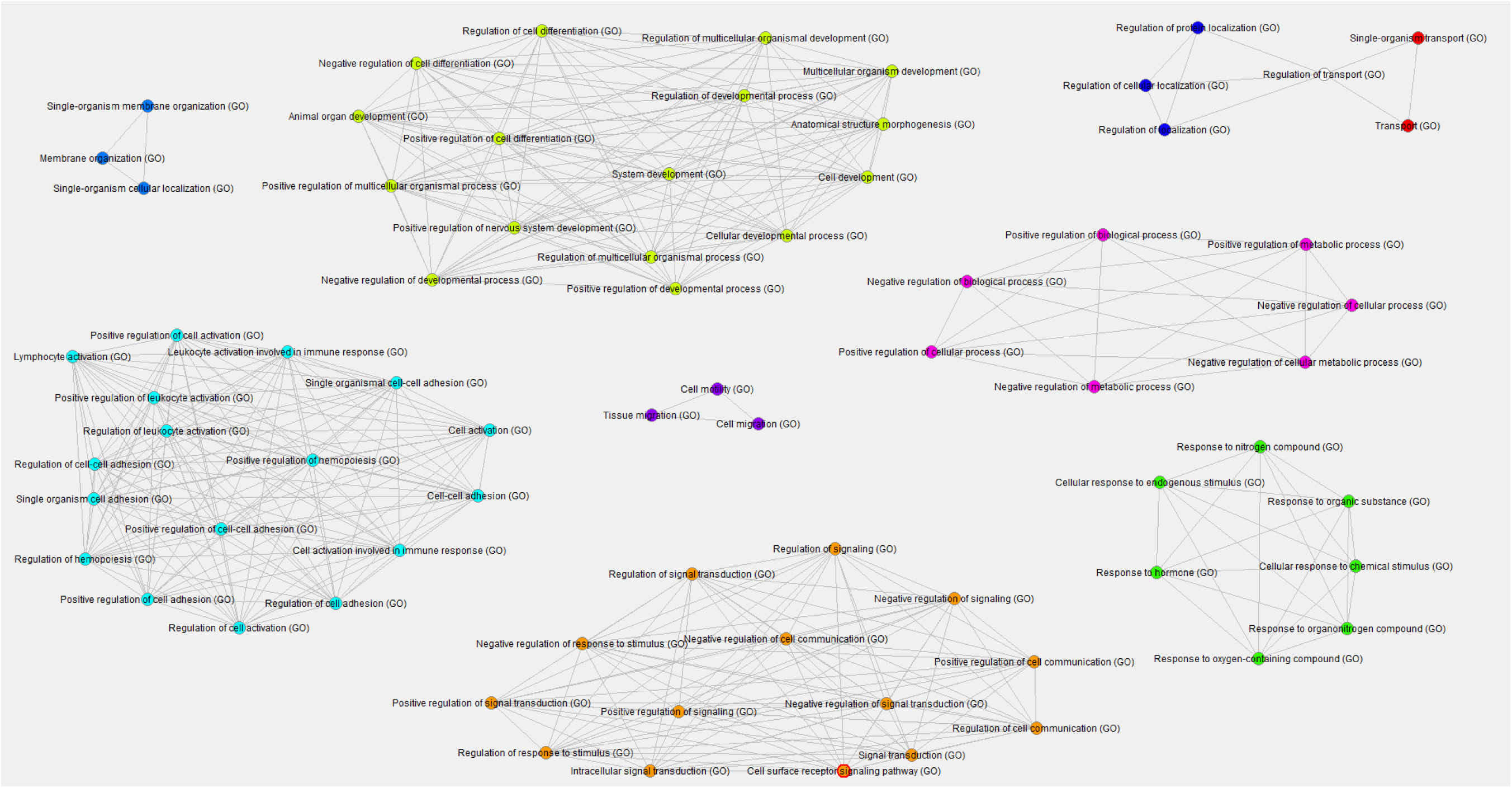
GO term network analysis. Biological process GO terms (level 3) clustered based on shared gene annotations. Nodes were only included if they belonged to a cluster of at least three terms and if they had an FDR < 0.05.

In FUMA analysis, we also observed enrichment in a variety of gene sets at FDR < 0.05. The largest number of gene sets was 145 immunologic signature gene sets (MsigDB c7; **Supplemental Table 8**), which represent cell states and perturbations within the immune system. For example, the top gene set in this collection was GSE40666 (UNTREATED-VS-IFNA-STIM-EFFECTOR-CD8-TCELL-90MIN-UP), which included 22 genes annotated to 29 CpGs, *P* = 8.8e-11.^63^ Other enriched gene sets (**Supplemental Table 9 to 16**) included 11 hallmark sets (MsigDB h), 71 chemical and genetic perturbation (MsigDB c2), 27 microRNA targets (MsigDB c3), 9 TF targets (MsigDB c3), 5 cancer gene modules (MsigDB c4), 4 oncogenic signatures (MsigDB c6), 13 WikiPathways, and 33 GWAS catalog reported genes.

### EWAS catalog analysis linked LE8-associated CpGs with a broad range of disease phenotypes

Among the 609 LE8-associated CpGs, 575 CpGs were reported to be associated with a variety of traits in the EWAS catalog.^17^ We identified 10 major phenotypes, including ischemic heart disease, stroke, chronic kidney disease, Alzheimer’s disease, chronic obstructive pulmonary disease, lung function (FEV1 and FEV1/FVC), chronic pain, rheumatoid arthritis, C-reactive protein, and pregnancy complications (hypertensive disorders of pregnancy, preeclampsia, and gestational diabetes mellitus). LE8-associated CpGs were enriched for all these 10 disease phenotypes (**Table 2**), e.g., 29-fold enrichment for stroke (*P* = 2.4e-15), 21-fold for ischemic heart disease (*P* = 7.4e-38), 22-fold for CKD (*P* = 1.3e-16), 6-fold for Alzheimer’s disease (*P* = 2.4e-6), and 7-fold for pregnancy complications (*P* = 1.5e-13). The phenotype with the largest number of overlapping CpGs was CRP; 562 of the 609 CpGs have been found to be associated with C-reactive protein (*P* = 1e-300). Lung function was the phenotype with largest fold enrichment in this analysis, 71-fold enrichment (*P* = 1.3e-14).

**Table 2:** Phenotype enrichment analysis in EWAS Catalog.

| Phenotype | Sub-Phenotypes | EWAS Catalog<br>CpG counts | Overlapping<br>CpGs | Fold<br>enrichment | +/- | Enrichment P |
| --- | --- | --- | --- | --- | --- | --- |
| Stroke | Incident Stroke & Self-Reported Prevalent Stroke | 339 | 13 | 28.8 | + | 2.4E-15 |
| Heart diseases | Incident Ischemic Heart Disease, Self-Reported Prevalent Ischemic Heart Disease | 1412 | 39 | 20.8 | + | 7.4E-38 |
| Lung function | FEV1, FEV1/FVC | 95 | 9 | 71.2 | + | 1.3E-14 |
| Chronic obstructive pulmonary disease | Incident COPD & Self-Reported Prevalent COPD | 38896 | 303 | 5.9 | + | 1.6E-155 |
| Chronic kidney disease | Chronic kidney disease, estimated glomerular filtration rate (eGFR), Incident Chronic Kidney Disease, urinary-albumin-creatinine ratio (UACR) | 557 | 16 | 21.6 | + | 1.3E-16 |
| Alzheimer's disease | Alzheimer's disease Braak stage, APOE e2 vs e4 | 1328 | 11 | 6.2 | + | 2.4E-06 |
| Pregnancy complications | Hypertensive disorders of pregnancy, Preeclampsia, gestational diabetes mellitus, very early preterm birth (<28 weeks) | 2504 | 24 | 7.2 | + | 1.5E-13 |
| Chronic pain | Incident Chronic Pain | 152 | 4 | 19.8 | + | 5.7E-05 |
| CRP | C-reactive protein | 64063 | 562 | 6.6 | + | 1.0E-300 |
| Rheumatoid Arthritis | Rheumatoid Arthritis & Prevalent Rheumatoid Arthritis | 52085 | 49 | 0.7 | - | 9.0E-03 |

We also searched for the association between LE8-associated CpGs and LE8 components in the EWAS catalog. We found that 452 CpGs have been associated with at least one of the six LE8 components (we found no overlap for physical activity associated CpGs and there was no study on sleep in the EWAS catalog). A Venn diagram for these 452 CpGs is provided in **Supplemental Figure 2**. The majority of these CpGs were associated with blood glucose (339 CpGs), smoking (213 CpGs), and/or BMI (116 CpGs) related phenotypes. We also found nine CpGs associated with diet, 16 CpGs with blood pressure, and 37 CpGs associated with blood lipids.

### PheWAS analysis highlights association of *cis*-meQTL SNPs with autoimmune diseases

Out of the 609 LE8 associated CpGs, 382 CpGs had at least one *cis*-meQTL SNP in the GoDMC database.^45^ We explored the associations between these SNPs and 1,358 phenotypes in the PheWAS of GWAS catalog SNPs.^44^ After Bonferroni correction (i.e., *P* < 0.05/1,358), we found 107 SNP-phenotype pairs (39 *cis*-meQTL SNPs for 11 CpGs; **Supplemental Figure 3**). In the 39 SNPs, 27 SNPs (*cis*-meQTLs for five CpGs) reside in the human leukocyte antigen (HLA) region of chromosome 6 (**Supplemental Table 17**). In addition, we observed that 31 SNPs (*cis*-meQTLs for eight CpGs) were linked with inflammatory or autoimmune disorders, e.g., gout, celiac disease, multiple sclerosis, type 1 diabetes, and rheumatoid arthritis (**Supplemental Table 18**). We further performed a PheWAS search for the 39 SNPs in the IEU OpenGWAS database; 35 SNPs were associated with at least one phenotype (range: 1 to 338; median: 13 phenotypes; **Supplemental Table 19**). Inflammatory or autoimmune disorders were also observed in the latter analysis. For example, consistent with the findings in the PheWAS catalog (**Supplemental Figure 4**), rs3135338 (*cis*-meQTL for cg00124375; annotated to *HLA-DRA*) was associated with type 1 diabetes (*P* = 1e-300),^64^ multiple sclerosis (*P* = 3.7e-294),^65^ and rheumatoid arthritis (*P* = 1e-200)^66^ in large GWAS.

### MR analysis suggests causal relationship of LE8-associated CpGs with multiple disease phenotypes

We performed two-sample MR analysis for the 382 CpGs with *cis*-meQTL SNPs and 14 phenotypes related to diseases identified by our EWAS catalog analysis. We observed 261 significant CpG-phenotype pairs for 141 unique CpGs at FDR < 0.05 (**Supplemental Table 20**). Overall, we found no strong evidence of horizontal pleiotropy or heterogeneity. Based on MR-Egger intercept, no CpG-phenotype pair had an intercept *P* < 1e-3. At *P* < 1e-3, heterogeneity was observed for 16 CpGs with four phenotypes (24 pairs). The majority of phenotypes are leukocyte composition (22 pairs). Nonetheless, leukocyte composition (% neutrophils, % monocytes, and % lymphocytes) were phenotypes with the largest numbers of significant CpGs; the number of associated CpGs were 57, 74, and 78, respectively (**Supplemental Table 21**).

We further examined the directions of associations from the LE8 EWAS and the MR analysis. That is, if the LE8 score was positively associated with DNAm levels at a CpG (i.e., hypermethylation), we expected that hypermethylation at this CpG was associated with a lower risk of disease phenotypes in the MR analysis, or vice versa. We highlighted 10 CpGs in **Table 3** (full significant MR findings for all CpGs are presented in **Supplemental Table 21)**. We showed four CpGs (of the 10 CpGs) with consistent associations in the two analyses. For example, we found that LE8 score was associated with hypomethylation at cg20544516 (*MIR33B*; *SREBF1; P* = 5.2e-10) and hypomethylation at this CpG was associated with lower risk of stroke (*P* = 8.1e-6). Similarly, we found that the LE8 score was associated with hypermethylation at cg26403580 (*PRDX3; P* = 4.8e-11) and hypermethylation at this CpG was associated with lower risk of stroke (*P* = 1.6e-4). However, we also observed inconsistent associations, e.g., the LE8 score was associated with hypomethylation at cg04379041 (*ZGPAT*; *P* = 2.0e-12) and hypomethylation at this CpG was associated with higher risk of heart failure (*P* = 6.2e-4).

**Table 3:**
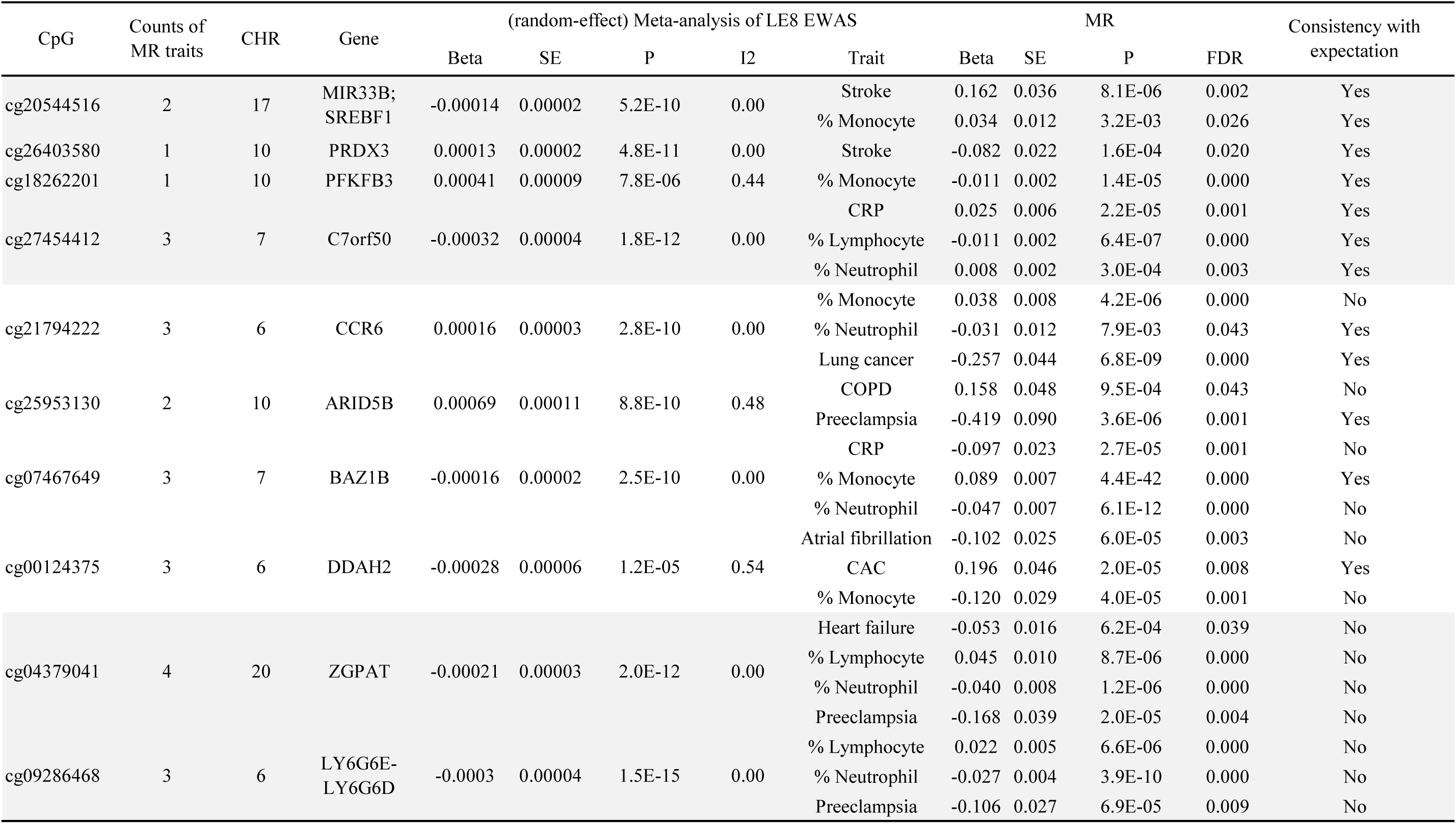
Selected MR analyses results for LE8-associated CpGs.

### CVH DNAm score predicts incident clinical outcomes

Of the 5,605 CpGs with FDR < 0.05 in the FHS, 1,326 CpGs were verified in CARDIA. Further, the elastic-net regression model selected 141 CpGs that were used to derive the CVH DNAm score in each cohort. This DNAm score was moderately associated with the LE8 score, e.g., Pearson *r* was 0.39 in FHS and 0.24 in MESA. The DNAm score by LE8 score deciles and density plot of the two scores in FHS and MESA are depicted in **Supplemental Figure 5**.

In FHS (**Table 4**), each standard deviation (SD) increase of the DNAm score was associated with striking risks of clinical outcomes. This included 34% (95% CI: 27%, 41%; *P* = 2.8e-14) lower risk of incident CVD, 29% (95% CI: 11%, 43%; *P* = 3.3e-3) lower CVD-specific mortality, and 35% (95% CI: 30%, 40%; *P* = 1.2e-16) lower all-cause mortality, adjusted for model 2 covariates. These associations were consistently observed in each of the other cohorts of diverse populations. The reduction in the risk of incident CVD, CVD mortality, and all-cause mortality per SD increase in the DNAm score ranged from 19% to 32%, 28% to 40%, and 27% to 45% (model 2 analyses), respectively. Compared to models with adjustment for sex, age, education, and leukocyte composition, adding the DNAm score increased the C statistic by average of 0.2 (**Supplemental Table 22**). The model 2 C statistic was 0.68, 0.84, and 0.79 in MESA and 0.72, 0.83, and 0.81 in JHS for incident CVD, CVD mortality, and all-cause mortality, respectively. Associations between the DNAm score and clinical outcomes in meta-analysis were similar with or without FHS are shown in **Figure 4**, without apparent cross-cohort heterogeneity.

**Figure 4:**
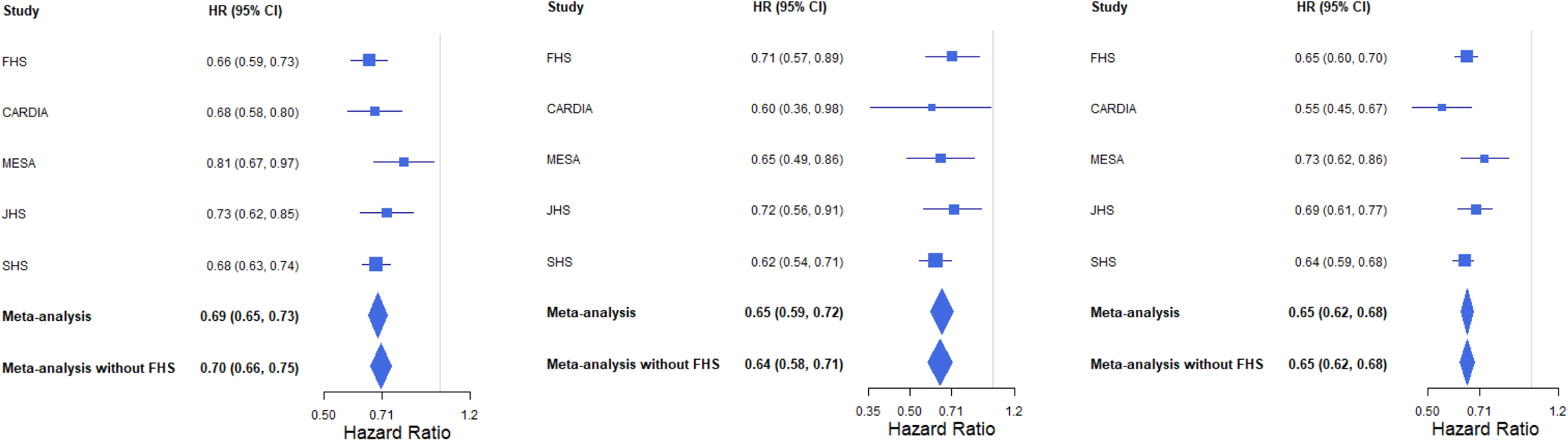
Hazard ratios for DNAm LE8 score for incident CVD, CVD mortality, and all-cause mortality. Hazard ratios are relative to one standard deviation increase in DNAm LE8 score. Model adjusted for sex, age, estimated leukocyte composition, cohort-specific covariates such as research centers, self-reported race/ethnicity, and education levels.

**Table 4:** Hazard ratios showing risk reduction for standard deviation increase in LE8 DNAm score.

| Cohort | N | Events | M1 |  |  | M2 |  |  |
| --- | --- | --- | --- | --- | --- | --- | --- | --- |
|  |  |  | HR | 95% CI | P | HR | 95% CI | P |
| Incident CVD |  |  |  |  |  |  |  |  |
| FHS | 3675 | 391 | 0.65 | (0.59, 0.72) | 2.9E-15 | 0.66 | (0.59, 0.73) | 2.8E-14 |
| CARDIA | 2609 | 168 | 0.65 | (0.56, 0.76) | 4.6E-08 | 0.68 | (0.58, 0.80) | 1.9E-06 |
| MESA | 926 | 125 | 0.81 | (0.68, 0.96) | 2.0E-02 | 0.81 | (0.67, 0.97) | 2.0E-02 |
| JHS | 1326 | 177 | 0.72 | (0.63, 0.84) | 1.4E-05 | 0.73 | (0.62, 0.85) | 6.7E-05 |
| SHS | 2064 | 901 | 0.67 | (0.24, 0.73) | 3.7E-21 | 0.68 | (0.24, 0.74) | 7.0E-19 |
| CVD Death |  |  |  |  |  |  |  |  |
| FHS | 3679 | 91 | 0.71 | (0.57, 0.89) | 3.2E-03 | 0.71 | (0.57, 0.89) | 3.3E-03 |
| CARDIA | 2609 | 18 | 0.58 | (0.37, 0.93) | 2.4E-02 | 0.6 | (0.36, 0.98) | 4.3E-02 |
| MESA | 926 | 53 | 0.63 | (0.48, 0.83) | 9.9E-04 | 0.65 | (0.49, 0.86) | 2.0E-03 |
| JHS | 1326 | 74 | 0.65 | (0.52, 0.81) | 1.1E-04 | 0.72 | (0.56, 0.91) | 6.9E-03 |
| SHS | 2064 | 399 | 0.6 | (0.21, 0.68) | 1.6E-15 | 0.62 | (0.22, 0.71) | 4.9E-13 |
| All-cause mortality |  |  |  |  |  |  |  |  |
| FHS | 4123 | 683 | 0.64 | (0.60, 0.69) | 1.0E-50 | 0.65 | (0.60, 0.70) | 1.0E-50 |
| CARDIA | 2632 | 108 | 0.54 | (0.44, 0.65) | 9.3E-11 | 0.55 | (0.45, 0.67) | 4.1E-09 |
| MESA | 926 | 159 | 0.71 | (0.61, 0.83) | 2.0E-05 | 0.73 | (0.62, 0.86) | 1.4E-04 |
| JHS | 1724 | 346 | 0.68 | (0.61, 0.75) | 2.8E-13 | 0.69 | (0.61, 0.77) | 3.6E-10 |
| SHS | 2064 | 1292 | 0.61 | (0.22, 0.66) | 8.8E-43 | 0.64 | (0.23, 0.68) | 3.5E-35 |

Covariates in M1 included sex, age, estimated leukocyte composition, cohort-specific covariates such as research centers, and self-reported race/ethnicity. Covariates in M2 included M1 covariates and education levels. Covariates in M3 included M2 covariates and LE8 score.

## DISCUSSION

In this new investigation including over 11,000 individuals with diverse ancestry and sociodemographic backgrounds, we identified an epigenetic signature comprising 609 CpGs consistently associated with the CVH as defined by the LE8 score. Enrichment analyses suggested these LE8-associated CpGs reflect a variety of specific biological pathways, particularly related to immune responses. EWAS catalog and PheWAS catalog analyses further demonstrated that these CpGs may be involved in the development of a range of interrelated diseases, showing the relevance of the health and behavioral factors that define good CVH in other disease pathways and overall health. Furthermore, utilizing genetic variants of the LE8-associated CpGs, we demonstrated that many of these CpGs were putatively causal for CVD phenotypes. Finally, we showed that an epigenetic CVH score could increase prediction for CVD, CVD-specific mortality, and all-cause mortality. Cumulatively, our study provides powerful new evidence that DNAm is a key mechanism to facilitate a better understanding of the underlying biology of CVD (and likely related conditions), informing future efforts for prevention and intervention of CVD.

Some earlier research has identified a limited number of DNAm sites associated with CVH. Primarily using CARDIA (n=1,085), Zheng et al. identified 45 CVH-associated CpGs.^15^ The present study builds on and greatly expands this prior work by evaluating the new AHA CVH metric, LE8, and substantially increasing the sample size and ancestry diversity of the population—ultimately identifying approximately ten times more CVH-associated CpGs. Importantly, the diversity of our study sample allowed us to identify CpGs shared across multiple ancestral populations. Beyond the advantage of guard against false positive signals due to minimizing population stratification-related confounding,^67, 68^ our multiethnic analysis increases both generalizability and public health relevance of our findings.

Our EWAS catalog analysis suggests that blood glucose, BMI, and smoking may be the major driving components for our LE8 EWAS findings. In addition, EWAS catalog enrichment, PheWAS, and MR analyses all showed that the LE8-associated CpGs are linked to several interrelated phenotypes. Although we did not directly examine the association between the LE8 score and all of these phenotypes, our observations suggest that LE8-associated phenotypes span more than just CVD. For example, we demonstrated enrichment of Alzheimer’s disease, which was consistent with recent studies showing that a higher LE8 score was associated with lower risk of dementia in UK Biobank.^69^ These findings highlight the foundational role of LE8 for promoting overall health.

One MR analysis finding regarding the association between hypomethylation at cg20544516 (*MIR33B*; *SREBF1*) and a lower risk of stroke was consistent with a recent study that found a relation of higher LE8 score to lower risk of stroke, supporting a putative causal relationship from LE8 to DNAm and then to stroke.^70^ To perform MR analyses, we used genetic variants that were identified by a large meQTL analysis in 32,851 individuals of European ancestry.^45^ This meQTL database, together the large GWAS databases for phenotypes of interest, provides a robust set of genetic variants to ensure the validity of our MR analysis. Nonetheless, we also observed some inconsistencies between our expectations and these empirical data, such as the association between hypomethylation at cg04379041 and increased risk of heart failure. One potential cause of bias may be reverse causality due to the cross-sectional design of the CVH EWAS. In this case, differential DNAm may lead to the onset of a disease phenotype, which then alters an individual’s CVH status, rather than CVH status leading to DNAm change and the disease phenotype. Our results support the need for future studies using longitudinal design with repeated DNAm measurements to provide additional evidence on the causal relationships between LE8, DNAm, and clinical outcomes.

Among the CpGs that were significant in the PheWAS analysis, many were annotated to genes that fall on the HLA region at chromosome 6.^52^ We showed the *cis*-meQTLs of these CpGs were associated with a variety of autoimmune diseases such as type 1 diabetes, rheumatoid arthritis, and celiac disease. CRP was the phenotype with the largest number of overlapping CpGs in EWAS catalog enrichment analysis, while leukocyte composition was the phenotype with the largest number of significant MR findings. These observations are consistent with prior studies reporting significant associations between LE8 components and inflammatory markers.^71-74^ Together with these prior reports, our analyses support a key role of immune response related mechanisms in mediating the relationship between LE8 and disease endpoints. Future experimental studies and clinical trials should consider and aim to confirm our observations.

This study included a diverse group of participants from six different cohorts, each well-established with rich data describing participants’ lifestyle, genetic, and clinical information. Although not all cohorts had complete data to calculate the LE8 score, we found high correlations of scores missing sleep or diet, as well as consistent results for the DNAm score and disease phenotypes across cohorts. While we evaluated six diverse cohorts with a range of ancestries, most were US-based, and future refinement and broader validation of our findings are needed in other world regions, including low-and middle-income nations where CVD is increasing most rapidly. Potential limitations should be considered. The behavioral components in LE8 scored were based on self-reported data using different instruments in different cohorts. Future use of more objective measurements of behaviors could further increase the strength of observed findings and associations. Heterogeneity in lifestyle and health factor assessment, as well as race/ethnicity, could also explain the relatively moderate EWAS replication rate (609 out of 5,223 CpGs). Better DNAm and LE8 data harmonization may improve statistical power for the remaining CpGs. DNAm data in the present EWAS was based on a single measurement; and repeated measures may reduce measurement error due to changes over time as well as allow assessment of temporal relationships. On the other hand, to-date empirical data on longitudinal change in DNAm is not well established.

Overall, we demonstrated that CVH, measured by LE8 score, was associated with strong DNAm signatures that can be observed across cohorts with diverse sociodemographic and ancestry backgrounds. Our study suggests that regulating immune response may be the key mechanism underpinning the association between LE8 and clinical outcomes. Future studies with large diverse study samples and longitudinal DNAm measurements are needed to better understand the causal relations between LE8 and DNAm, as well as their clinical consequences.

## Disclaimer

The views and opinions expressed in this manuscript are those of the authors and do not necessarily represent the views of the National Heart, Lung, and Blood Institute, the National Institutes of Health, or the U.S. Department of Health and Human Services.

## Data availability

The datasets analyzed in the present study are available at the dbGAP repository phs000285.v3.p2 for CARDIA (https://www.ncbi.nlm.nih.gov/projects/gap/cgi-bin/study.cgi?study_id=phs000285.v3.p2), phs000007.v33.p14 for FHS (https://www.ncbi.nlm.nih.gov/projects/gap/cgi-bin/study.cgi?study_id=phs000007.v33.p14), phs000209.v13.p3 for MESA (https://www.ncbi.nlm.nih.gov/projects/gap/cgi-bin/study.cgi?study_id=phs000209.v13.p3), phs000286.v7.p2 for JHS (https://www.ncbi.nlm.nih.gov/projects/gap/cgi-bin/study.cgi?study_id=phs000286.v7.p2), phs000580.v1.p1 (https://www.ncbi.nlm.nih.gov/projects/gap/cgi-bin/study.cgi?study_id=phs000580.v1.p1).

## Data Availability

The datasets analyzed in the present study are available at the dbGAP repository.

